# Pushing the limits of recovery in chronic stroke survivors: User perceptions of the Queen Square Upper Limb Neurorehabilitation Programme

**DOI:** 10.1101/2020.01.05.19015099

**Authors:** K Kelly, F Brander, A Strawson, NS Ward, KS Hayward

**Affiliations:** The National Hospital for Neurology and Neurosurgery, Queen Square, London WC1N 3BG, United Kingdom; Department of Clinical and Motor Neuroscience, UCL Queen Square Institute of Neurology, Queen Square, London WC1N 3BG, United Kingdom; Department of Physiotherapy, Melbourne School of Health Sciences; AVERT Early Rehabilitation Research Group, Stroke Theme, Florey Institute of Neuroscience and Mental Health; NHMRC Centre of Research Excellence in Stroke Rehabilitation and Brain Recovery; University of Melbourne, Heidelberg VIC 3084, Australia

**Author notes:** Corresponding author: Dr Kathryn Hayward. Department of Physiotherapy, Melbourne School of Health Sciences; AVERT Early Rehabilitation Research Group, Stroke Theme, Florey Institute of Neuroscience and Mental Health; NHMRC Centre of Research Excellence in Stroke Rehabilitation and Brain Recovery; University of Melbourne, Heidelberg VIC 3084, Australia. equal first author (K Kelly, F Brander).

**Keywords:** stroke, upper limb, rehabilitation, qualitative, focus groups

## Abstract

**Introduction:** The Queen Square Upper Limb (QSUL) Neurorehabilitation Programme is a clinical service within the National Health Service in the United Kingdom that provides 90 hours of therapy over three weeks to stroke survivors with persistent upper limb impairment. This study aimed to explore the perceptions of participants of this programme, including clinicians, stroke survivors and carers.

**Design:** Descriptive qualitative.

**Setting:** Clinical outpatient neurorehabilitation service.

**Participants:** Clinicians (physiotherapists, occupational therapists, rehabilitation assistants) involved in the delivery of the QSUL Programme, as well as stroke survivors and carers who had participated in the programme were purposively sampled. Each focus group followed a series of semi-structured, open questions that were tailored to the clinical or stroke group. One independent researcher facilitated all focus groups, which were audio-recorded, transcribed verbatim and analysed by four researchers using a thematic approach to identify main themes.

**Results:** Four focus groups were completed: three including stroke survivors (n = 16) and carers (n =2), and one including clinicians (n = 11). The main stroke survivor themes related to *psychosocial* aspects of the programme (“you feel valued as an individual”), as well as the *behavioural training* provided (“gruelling, yet rewarding”). The main clinician themes also included *psychosocial* aspects of the programme (“patient driven ethos – no barriers, no rules”), and *knowledge, skills and resources* of clinicians (“it is more than intensity, it is complex”).

**Conclusions:** As an intervention, the QSUL Programme is both comprehensive and complex. The impact of participation in the programme spans psychosocial and behavioural domains from the perspectives of both the stroke survivor and clinician.

**Strengths and limitations:**

- Descriptive qualitative study of the perception of users (stroke survivor, carer, clinician) engaged in the delivery of the Queen Square Upper Limb Neurorehabilitation Programme, which is run at a single centre in the United Kingdom.
- Focus groups were completed by a researcher independent of the programme, without the involvement of senior management to facilitate open discussion and critical reflection of the programme.
- This study involved a sample of users that were involved in the programme in the previous 12-months.
- Data coding was performed by four researchers enhancing the validity of the results.
- As only two carers were included in the focus groups, their experience has limited representation in the results.

## Introduction

The burden of upper limb impairment after stroke remains high with up to 70% of survivors experiencing persistent difficulty using their affected upper limb six months post-stroke (1-3). Recent clinical trials of treatments targeting the post-stroke upper limb have been underwhelming, possibly because the content of the treatment was ineffective or the dose was too low (13-36 hours total or ∼30minutes per day) (4-6). Two related trials in which 300 hours of upper limb therapy was delivered to chronic (>6-months) stroke patients over 12-weeks were however strikingly positive (7, 8). In line with this, several meta-analyses suggest that larger doses of therapy (>2hrs per day) lead to clinically meaningful improvements of the upper limb post-stroke (9, 10). The effective dose (measured in time) of neurorehabilitation is likely to be much higher than that tested in recent clinical trials.

The Queen Square Upper Limb (QSUL) Neurorehabilitation Programme provides high dose, high intensity (90 hours over three weeks) neurorehabilitation for (mostly) chronic stroke survivors (11). The results are clinically meaningful and quantitatively similar to those reported by McCabe et al (7) and Daly et al. (8). Given high dose therapy is provided, the QSUL Programme presents a unique opportunity to investigate the components of successful post-stroke upper limb neurorehabilitation and include the users’ perspective. Therefore, the aim of this study was to explore (i) stroke survivor (and carer) and (ii) clinician (physiotherapist, occupational therapist, rehabilitation assistant) perceptions of the key therapeutic ingredients of post-stroke upper limb neurorehabilitation.

## Methods

### Overview of design and methods

A descriptive qualitative design was used. Focus groups with semi-structured, open questions and prompts were used to collect group perceptions and experiences. This study was registered with University College London Hospitals (UCLH) National Health Service Trust clinical audit and service development department as a service evaluation (See appendix 1 for details). All participants provided written informed consent to participate and for voice recording.

A purposive sample of London-based stroke survivors (and carers if available) who had completed the QSUL Programme within the previous 12-months were invited (face-to-face, telephone or email) to attend one of three focus groups. Potential participants were selected by management and senior clinical staff working on the programme (KK, FB, AS) with the view to include those from whom most could be learnt and to ensure a wide range of opinions were captured. Sample variation in sex, age, time post-stroke and stage of programme follow-up were targeted. All clinicians who had been or were currently involved in delivery of the programme were invited to participate, including physiotherapists, occupational therapists and rehabilitation assistants. The managing clinical team members (KK, FB, NW) of the programme were not invited to participate to facilitate free sharing of perceptions by clinicians.

### Data collection

Focus groups were performed in a quiet room within the hospital using a semi-structured question guide (Box 1), which included main questions and prompts. The facilitator (KH, PhD, Research Fellow) was independent of the QSUL Programme, had not been part of any participant therapy or assessments, and was not part of the managing clinical team. Her role was to encourage participants to share their personal experiences and opinions as to the key components of the programme (as a stroke survivor, carer or clinician) and used probing techniques and prompts to achieve further in-depth reflection. At the end of the focus group, the facilitator rephrased main experiences and meanings expressed to ensure accurate interpretation of participant views. Each focus group was audio-recorded and an additional independent person (final year physiotherapy placement student) took field notes during each focus group. The facilitator and field note personnel discussed each focus group at its end to corroborate main discussion points and notes.

#### Box 1

Focus group guide, including main questions and prompts

| **Patients and carers** | **Therapists** |
| --- | --- |
| Q1: Tell me about your experience of being involved in the day-to-day delivery of the QSUL Programme.<br>Probes:<br>How does this training programme differ to others?<br>What are your thoughts on schedule?<br>What are some positive experiences?<br>What are some negative or challenging experiences?<br>Q2: How does this training programme impact recovery of upper limb function (impairment/ activity/participation) post-stroke?<br>Probes:<br>What about the training positively influenced recovery?<br>What about the training negatively influenced recovery?<br>Have you found ways to get around/overcome these?<br>How could the programme be improved? | Q1: Tell me about your experience of being involved in the day-to-day delivery of the QSUL Programme.<br>Probes:<br>How does this training programme differ to others?<br>What are some positive experiences?<br>What are some negative or challenging experiences?<br>Who do you think benefits most from the programme? Why?<br>Q2: How does this training programme impact recovery of upper limb function (impairment/ activity/participation) post-stroke?<br>Probes:<br>What about the training positively influences recovery?<br>What about the training negatively influences recovery?<br>Have you found ways to get around/overcome these?<br>How could the programme be improved? |
| Q3: What are the active ingredients of the programme?<br>Probes:<br>What aspects of the programme are essential?<br>What aspects are not essential/lower priority? Why?<br>What about therapist skill set? Additional staffing<br>What about environment/resources/living/travel supports etc? | Q3: What are the active ingredients of the programme?<br>Probes:<br>What aspects of the programme are essential?<br>What aspects are not essential/lower priority? Why?<br>What about therapist skill set? Additional staffing<br>What about environment/resources/living/travel supports etc? |

### QSUL Programme

For a full description of the programme including staffing levels please see Ward et al., (11). In brief, the QSUL Programme provides 90 hours of therapy over three weeks, with follow-up in an outpatient clinic at six weeks and six months post programme completion. Two patients are admitted to the programme each week as day attendees (six patients are on the programme at one time), either from home or University College London Hospital dedicated patient hotel if they are self-caring, or self-caring with the support of one person. Daily intervention consists of six-hours of scheduled therapy including two sessions each of one-on-one occupational therapy and physiotherapy focused on analysis of movement and tasks, reduction of impairment and re-education of motor control within functional tasks. This is supplemented with two sessions of tailored individualised therapy with a rehabilitation assistant targeting repetitive task practice, sensory retraining, adjuncts to therapy such as functional splints, neuromuscular electrical stimulation, robotic devices and group work. Furthermore, patients are encouraged to work independently on cardiovascular fitness and are provided with homework to complete each weekend. Education, goal setting and developing self-efficacy for recovery are integral components that occur throughout the programme.

### Data analysis

Baseline clinical measures were collected during the programme (see (11)) and participant demographics were confirmed prior to the start of each focus group. Verbatim transcription was performed by a professional transcription agency (K-International, UK). A thematic content approach was used (12). Four researchers performed data analysis to avoid any potential bias or personal motivations, promoting confirmability. First, researchers (KK, FB, AS, KH) independently read and became familiar with the complete data set. Second, researchers went through the transcripts line by line to obtain meaningful information and identify repeated topics and patterns. Researchers then interactively discussed interpretation of data to avoid bias in analysis, and integrated data into themes. Credibility was enhanced through repeated discussions during the analysis process to (i) clarify interpretation of the data, (ii) reframe key themes and subthemes confirming consistency of findings between researchers, and (iii) ensure that defined themes accurately reflected the expressions of the participants. Next, quotations and sections of text were extracted under thematic content and checked for consistency with the narrative theme. Finally, on two occasions two researchers (AS, KH) re-read all transcripts to confirm that all data fitted into the identified themes and subthemes: post completion of theme development and post manuscript write-up. During the writing stage, further refinement of links and subthemes occurred to ensure consistency of themes. All changes were discussed at each step between the four researchers to achieve consensus. Final transcripts and results of the analysis were not discussed with participants.

## Results

### Stroke patients and carers

From 39 stroke survivors invited to participate, two declined, 17 were unable to attend and four did not attend as planned. Sixteen stroke survivors (8 male, 8 female) and two carers (1 male, 1 female) participated in one of three focus groups. See Table 1 for stroke participant characteristics. The mean focus group duration was 79.7 minutes.

**Table 1:** Demographics of stroke survivors, n = 16

| Characteristic |  |
| --- | --- |
| **Age, years, median (IQR)** | 58 (48 – 69.3) |
| **Sex** |  |
| Female, n (%) | 8 (50) |
| Male, n (%) | 8 (50) |
| **Months since stroke, median (IQR)** | 19 (12.5 – 30.3) |
| **Modified Fugl-Meyer Upper Limb, /54, median (IQR)** | 35 (23 – 43.5) |
| **Paretic upper limb** |  |
| Left, n (%) | 12 (75) |
| Right, n (%) | 4 (25) |
| **Proportion dominant upper limb affected** |  |
| Dominant, n (%) | 7 (43.75) |
| Non-dominant, n (%) | 9 (56.25) |
| **Family support available, self-reported, n (%)** | 15 (93.8) |
| **Modes of QSUL programme access** |  |
| Taxi vouchers, n (%) | 5 (31.3) |
| Hotel accommodation, n (%) | 10 (62.5) |
| Underground train, n (%) | 1 (6.3) |
| **Employment status at QSUL programme enrolment** |  |
| Student, n (%) | 2 (12.5) |
| Retired, n (%) | 5 (31.3) |
| Not working, n (%) | 4 (25) |
| Working, n (%) | 4 (25) |
| Volunteering, n (%) | 1 (6.3) |

### Overview of themes for stroke survivors and carers

Two main themes, each containing subthemes, were identified from the transcripts and are presented in Table 2.

**Table 2:**
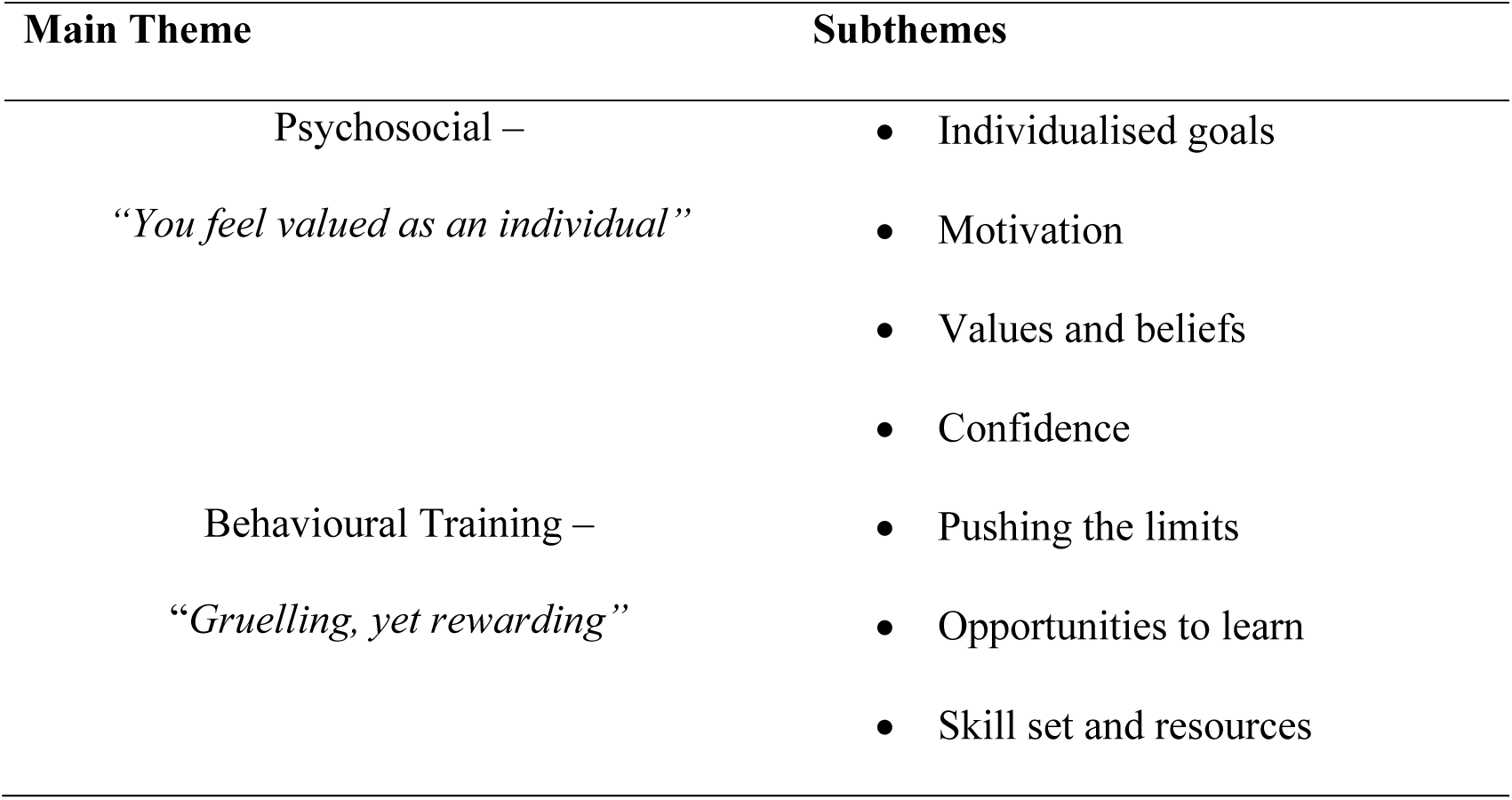
Summary of themes identified from stroke survivor and carer focus groups.

Theme 1 Psychosocial: “You feel valued as an individual”

Stroke survivors and their carers consistently discussed the psychosocial components of the programme. Four subthemes were constructed from the data that were critical to enhancing participation during the three-week programme, as well as maintaining motivation for recovery over the following six months of programme follow-up.

### Individualised Goals

Stroke survivors identified that the programme gave them the opportunity to set personalised goals collaboratively with an occupational therapist and physiotherapist, which impacted on their relationship with clinicians and engagement in the programme.

*‘Here’s your thing - this is individualised, tailored to you, your needs, your goal*.*’*

Stroke survivors frequently discussed that they were encouraged to set ambitious and challenging goals with nothing considered off limits.

*‘You know you’re going to take it in stages, you’re going to build up to it but there’s nothing – you don’t get a no*.*’*

Stroke survivors highlighted that while most goals were focused on their upper limb, they were encouraged to define goals related to their daily routine and/or leisure interests. A broader scope for defining goals meant that stroke survivors had the opportunity to experience the benefits of using their arm and hand more often in everyday situations.

*‘It was so good that you could bring in like your home life experiences, so it wasn’t just doing things*.*’*

### Motivation

Stroke survivors discussed how motivation to persist with the programme was drawn from a variety of sources. This included the enriched rehabilitation environment, variability of activities and incremental task progression throughout the programme. Additionally, the focus on meaningful real-world tasks was considered important to improve intrinsic motivation, and to maintain interest in working towards upper limb recovery.

*‘I found doing exercises all day very boring and difficult and not very motivating but when you’re doing stuff that’s actually fun and stimulating like playing tennis – it’s doing the same stuff but you’re having fun*.*’*

All stroke survivors discussed the high levels of support received throughout the programme, from both clinicians and fellow stroke survivors who may have had similar problems currently or in the past. The collaborative team focus of the programme, where stroke survivors and clinical staff are working in the same space, provided opportunities for enhanced motivation and self-efficacy; driven by observation-in-action.

*‘I didn’t find any inhibitions when I came here. I didn’ t feel embarrassed once I was here with … with our little group*.*’*

*‘When you work with a group you get some motivation as well because when you see the other people, they can do it, I can do it*.*’*

The impact of intrinsic motivation on achieved outcomes and recovery was discussed. Stroke survivors described observing some patients on the programme who had a lack of the ‘right attitude’, which was perceived to hinder recovery and potentially limit derived benefit from the programme.

*‘It’s only going to work if you come with a positive attitude and you believe in yourself*.*’*

*‘I think after a while you adapt to your limitations and you don’ t try*.*’*

The structure of the programme along with the follow-up appointments was described as integral to carry-over into the home environment. Knowing they were coming back for a follow-up appointment was considered to increase drive to continue with therapy after completing the programme.

*‘But you want to do it for them too because they’ve put a lot of work into helping you and you don’t want to let them down*.*’*

### Values and Beliefs

Many of the participants reported feeling quite negative regarding their rehabilitation potential on discharge from previous therapy services or programmes. This created anihilistic attitude towards recovery, as many patients were led to believe that they could not influence or drive their own progress.

*‘I was written off the year before but here it was “no, we can work on that” …’*

The positive attitude of clinicians on the QSUL Programme was described as essential to help each individual acknowledge that they had the potential to improve their recovery, independently participate in the community, and ultimately take ownership of their rehabilitation.

*‘…Puts you on the road – what to do to help yourself*.*’*

### Confidence

All of the above subthemes were reported to have an extremely positive effect on the stroke survivors’ confidence in their daily routine and activities, creating a sense of autonomy. *‘After the three-week programme I got some confidence for myself and now I can go around … getting involved in all activities, social-media and everything … So it’s both mentally and physically helpful*.*’*

Participants highlighted that being removed from their home environment, and their habitual routine and supportive families further enhanced their confidence in their own ability to be independent. Those that required use of programme access enablers e.g., using taxis or staying in the hotel, described these to positively influence independence and in turn, confidence.

*‘Managing my timing and getting up and sorting and getting to the breakfast room gave me a lot of confidence*.*’*

*‘I came here by myself – on the train and taxis and things like that, for me that was a big thing*.*’*

The well organised, positive team approach was considered important for building confidence for success. The opportunity to successfully achieve their goals by practice and repetition of tasks with feedback also contributed to confidence building.

*‘It’s also what they offered, the team. The team made you feel positive and made you feel things were achievable*.*’*

Theme 2: Behavioural Training: “Gruelling, yet rewarding”

The programme was described to provide the opportunity for stroke survivors to participate in gruelling, high-intensity behavioural training, which all stroke survivors reported that they relished. On completion of the programme, stroke survivors described that they had greater understanding of what their capabilities were, what was possible for the future and how to progress their training.

### Pushing the Limits

All stroke survivors acknowledged that the programme was exhausting, but the benefits of the intensity were superior. Some stroke survivors reported fatigue at the end of each day. Only one participant reported that it interfered with participation in the programme, which was able to be accommodated within the flexible structure of the programme.

*‘The harder they (clinicians) push the better the result and whilst it’s gruelling, those that are happy to accept it … will know at the end of the day that they are going to be better*.*’*

*‘I mean I was that completely shattered and … by the end of the week I couldn’t wait to stop the programme … I must confess, but I knew it was so, so helpful*.*’*

All stroke survivors agreed that having a structured timetable whilst on the programme was useful, giving them something to stick to, even when they may have felt like stopping. They felt the timetabling was tailored to the needs of the individual and was important to maintain a focus on therapy time, providing intensity and repetition of practice with variety.

*‘The structure forces you to extend yourself and you need to be forced*.*’ ‘I think it’s important to keep a timetable. You can’ t slack off*.*’*

Critical to being able to push the limits were access enablers. For example, the close location of the hotel was considered by stroke survivors to minimise fatigue, and enable longer duration of active participation in the intense programme as distance barriers were removed.

### Opportunities to Learn

Tackling activities that were not able to be performed prior to attending the programme was important to participants. Trialling of new ideas to solve old problems was a unique experience, from which they learnt how to engage in behavioural training and real-world practice. Some participants described that the problem-solving skills and knowledge which they learnt on the programme had been carried over to help them solve new tasks when returning home. Many found the holistic approach and integration of physiotherapy and occupational therapy useful to learn new skills for overall recovery.

*‘If we couldn’t do something one-way an alternative way was shown. I think it really came together because you could see how much you were learning*.*’*

The opportunity to access gym equipment and aids for activities of daily living provided greater variety, as well as specificity within individual behavioural training. Stroke survivors reported the positive impact of extension of rehabilitation opportunities into the community when linked to their goals, for example access to pushbikes, local gyms or swimming pools.

*‘Using the equipment, because I didn’t think I would even try any equipment, but after I finished here I went and joined the local gym*.*’*

*‘They took me swimming and I played tennis and those were the two big things*.*’*

### Skill Set and Resources

The stroke survivors stressed the importance of the skillset and expertise of the clinicians on the programme, as well as the collaborative relationships between clinician-patient and physiotherapist-occupational therapist. The importance of integration of all skillsets and communication between all team members when delivering the service was considered to have a marked effect on the success of the programme.

*‘The fact that the course [programme] is physio and OT was so desperately important. ‘‘I think it was a team effort …and between the two, the OT and the physiotherapist that you knew exactly what was going on*.*’*

The skillset and creativity of the clinicians was considered essential to breakdown goals into achievable components, adapt techniques and adjust treatment modalities to allow goal practice.

*‘Therapists would say: ‘Let’s try a different way of either working with the problem or looking at the problem*.*’*

Stroke survivors perceived that small group sizes and a well-resourced environment was beneficial in supporting clinicians and important in programme success.

*‘Well they keep the groups fairly small … so they’ve actually got the time*… *so they can see each individual*.*’*

*‘You can’t do the same thing unless you resource it in the same way*.*’*

## Clinicians

Eleven clinicians (1 male, 10 female) participated in one focus group. One invited clinician was unavailable at the time of the focus group (Band 7 physiotherapist). See Table 3 for characteristics of included clinicians. The focus group duration was 73.1 minutes.

**Table 3:** Demographics of clinicians, n = 11

| Characteristics |  |
| --- | --- |
| **Gender** |  |
| *Female, n (%)* | 10 (91) |
| *Male, n (%)* | 1 (9) |
| **Clinical profession** |  |
| *Occupational Therapist, n (%)* | 4 (36.4) |
| *Physiotherapist, n (%)* | 5 (45.5) |
| *Rehabilitation Assistant, n (%)* | 2 (18.2) |
| **Banding and years of clinical practice** |  |
| *Band 7 (Highly Specialist Therapist), n (%), average years of practice* | 6 (54.5), 11 |
| *Band 6 (Specialist Therapist), n (%), average years of practice* | 3 (27.3), 6.3 |
| *Band 3 (Rehabilitation Assistant) n (%), average years of practice* | 2 (18.2), 1.2 |

### Overview of themes for clinicians

Two main themes, each containing subthemes, were identified and are presented in Table 4.

**Table 4:**
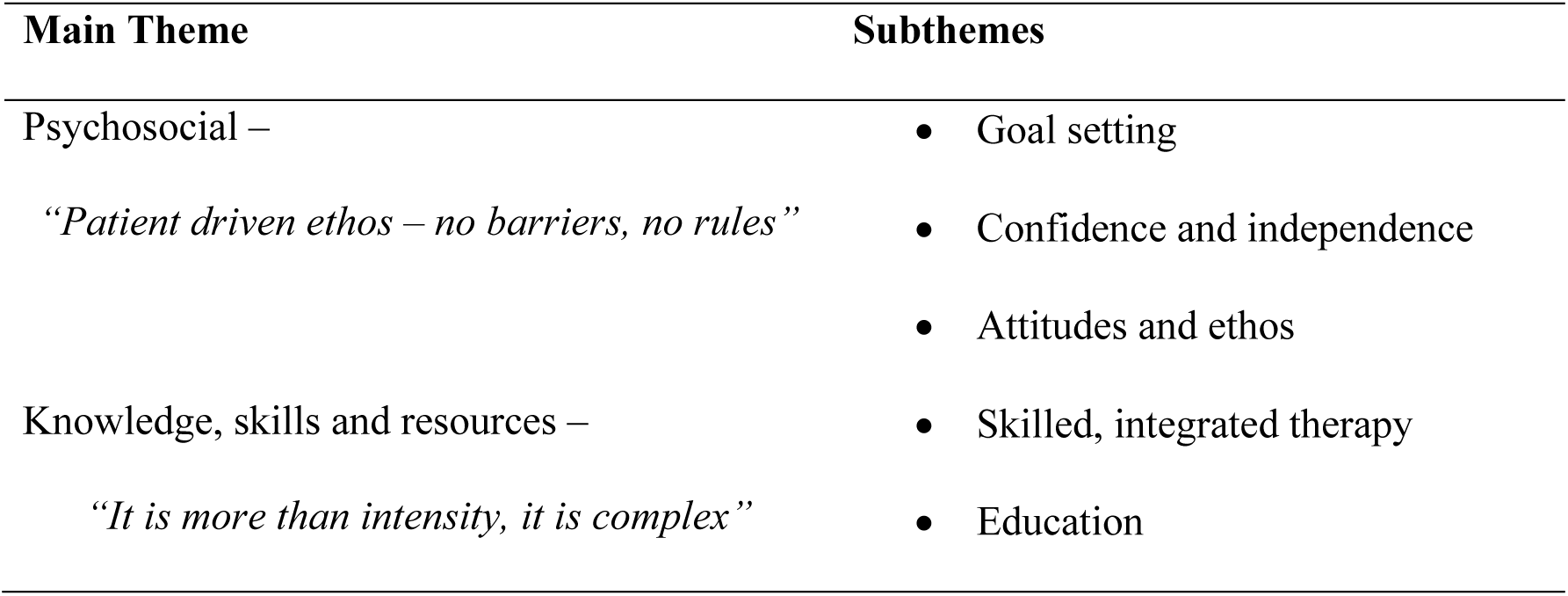
Summary of themes identified from clinician focus group.

Theme 1: Psychosocial

### Goal Setting

Clinicians highlighted the importance of individualised collaborative goal setting with stroke survivors. Some mentioned the difficulties of setting functional goals when stroke survivors had very little movement and/or had achieved little recovery to date. Within the focus group, clinicians highlighted that they had the time to access a variety of resources as useful tools for developing stroke survivor engagement in their recovery and goal attainment.

*‘We have the time and the resources to focus thoroughly on what is a really key player in daily life and we all know that the upper limb is missed…’*

A strategy described by many clinicians to support goal-achievement was education about functional task practice or activities rather than impairment-based goals. Previous clinical experience and knowledge of goal setting processes was considered essential.

*‘Because we give them license to be quite creative and to be a bit aspirational and you can actually do goals that are really, really specific to the patient*.*’*

### Confidence and Independence

The clinicians acknowledged that some of the gains made by stroke survivors during the programme related to improved confidence; not only in the ability to use their arm in tasks, but also in trying new tasks or skills, and persevering if they were not immediately successful. Clinicians perceived that stroke survivors also became more confident to participate in community tasks, leisure interests and in their ability to look after themselves, enhancing self-worth and identity.

*‘Surprised at how low and little confidence patients have. Also, how that confidence increases during the three weeks. I think it’s a nice safe place for patients to try out different things*.*’*

*‘I think it’s good for some of the patients that aren’t given much independence at home when their partners are not with them and they are encouraged to be more independent and do things by themselves*.*’*

In addition, clinicians highlighted the support amongst the stroke survivors. Each group of stroke survivors became close-knit, encouraging and motivating each other during the programme, aiding with confidence building

*‘I think there’s a lot of camaraderie between them as well. I think they get a lot from each other and I think being staggered helps people to see each other on their own journey*.*’*

### Attitudes and Ethos

Clinicians described the burden of high expectations from stroke survivors and programme management to deliver an intense programme with successful outcomes.

*‘I mean they’ve been told in clinic what they’ll be getting and they’ve been told they’ve got potential to get better*.*’*

Clinicians felt the ethos of the programme promoted a very open culture, allowing time and freedom to be creative around therapeutic and behavioural interventions. Many clinicians felt that anything was permitted on the programme and there were no barriers or rules to be broken.

*‘The whole culture and attitude is very much there’s nothing you can’t do which I think is really lovely*.*’*

*‘I have never been able to take patients swimming, patients to a tennis court before*.*’*

Clinicians acknowledged that there might be a positive bias in terms of the type of patient on the programme, in that stroke survivors had often actively sought referral to the clinic meaning that on the whole they had a drive to improve and willingness to learn. Clinicians also suggested that stroke survivors have to buy into the ethos of the programme, understanding and subscribing into the recovery process in order for it to be effective.

*‘I kind of feel like you’ve got them at the right time and the people that are being referred are clearly the ones seeking out further rehab so they are the ones that are motivated*.*’*

The clinicians highlighted that there was a subset of stroke survivors that required more support and demonstrated increased reliance on clinicians, with less understanding and buy-in to the self-management aspect of the programme.

*‘The ones that get the most out of the programme are the ones that do take up the independent time to carry on something you’ve set up. More chance of carrying it over*.*’*

Clinicians identified that stroke survivors’ outcomes from the programme were not just due to intensity of hands-on therapy, behavioural training and ability to build on training day after day, but rather the ethos of the programme along with the holistic, integrated approach and multidisciplinary nature of the programme. It was more than just repetitions of movements. *‘It’s not intensity by itself, it’s intensity of everything we’ve talked about –impairment, function, education, goals, independent tasks, homework, groups…*.*’*

Theme 2: Knowledge and Skills

### Skilled, integrated therapy

An important aspect of the programme that enabled smooth running was the skill and level of staffing. It was emphasised that the skillset was integral to implement a structured, yet flexible timetable to meet the varied needs of each stroke survivor. Many clinicians thought it was crucial to have previous neurological rehabilitation experience if you were to be a clinician on the programme, resulting in highly skilled clinical expertise and reasoning.

*‘The therapist’s skill and knowledge base is the really crucial thing – like handling and creative problem solving*.*’*

The clinicians understood each discipline’s unique skillset and role, which enhanced their ability to work collaboratively. They highlighted the interdisciplinary working and holistic approach and the impact this had on stroke survivor outcomes. Teamwork and open communication were identified as essential to enable clinicians to learn from and support each other, enhancing their own skillset.

*‘It’s not just physios working on impairment and OT’s looking at function, it’s really integrated which is really good*.*’*

### Education

A key component of the programme described was education, both for the stroke survivors and their carers. The clinicians identified that a significant amount of time was spent throughout the programme educating stroke survivors about stroke, their upper limb and how to improve. This was done through impairment-based training, retraining quality of movement whilst performing daily activities and practising goals in real-world environments.

*‘You’re not getting people in three weeks to change there and then but it’s getting them to see how change can happen for years and years. Education is the foundation, so they go home and continue*.*’*

Education was also described as useful to overcome barriers to buy-in. Some were described as more difficult to overcome including fatigue, cognitive deficits and negative health beliefs.

*‘I don’t think they always come in with the best…. understanding of why they can’t use their arm so there’s a lot of – we talk a lot during sessions*.*’*

## Discussion

In developing and implementing an intervention, especially one that is complex, it is crucial to understand and define the key components involved. Here, we report the stroke survivor, carer and clinician perceptions of the key components of the QSUL Programme. Not surprisingly there were overlapping themes; both the stroke survivor/carer and clinician groups considered the psychosocial aspects of the programme equally important as the behavioural training and intensity. Psychosocial and behavioural training includes individualised goal setting; building confidence and independence; attitudes and ethos; skilled, integrated therapy; and education. The skilled, integrated therapeutic approach enabled experimentation of practice in order to achieve success in their individualised goals, sustaining motivation (13), improving confidence and self-efficacy (14). Both groups considered education as vital to enable stroke survivors to drive their own recovery beyond the structured clinical rehabilitation environment.

This study suggests that successful post-stroke neurorehabilitation is likely to be a complex intervention (15). Complex upper limb therapy interventions (5, 8), including the current programme (11), have between eight to 12 key components/ingredients (see Table 5a). In contrast, the most commonly investigated intervention approaches, including constraint-induced movement therapy, repetitive task training and robotics, include far fewer components (see Table 5b). Constraint induced movement therapy does include behavioural (intensive therapy) and psychosocial methods (to enhance transfer to real-world environments). Of the complex intervention approaches tested, only iCARE was an adequately powered, multicentre, randomised controlled trial (5). The iCARE trial, however, had a neutral outcome as the findings were likely confounded by the low dose (30 hours over 10 weeks). Consistent with the QSUL Programme, the iCARE intervention did have a focus on the psychosocial aspects of behavioural training including active patient involvement in goals and development of self-management skills. McCabe et al., (7) and Daly et al., (8) were both before-after clinical studies that demonstrated large, clinically meaningful results from an intensive training protocol (300 hours over 12 weeks) that included the most behavioural training elements. They had little focus on the psychosocial components such as individualised goal setting and education, which were highlighted in this current study as key components. All three complex interventions did have an emphasis on treating at an impairment level and using meaningful functional activities to encourage participation, with a focus on quality of movement. If we move to accept the premise that effective rehabilitation is complex and multicomponent, then we are unlikely to work out the optimal combination of active ingredients simply by studying each in isolation. Understanding the combinations of ingredients that span effective complex interventions is thus critical to guide the next clinical trials.

**Table 5:**
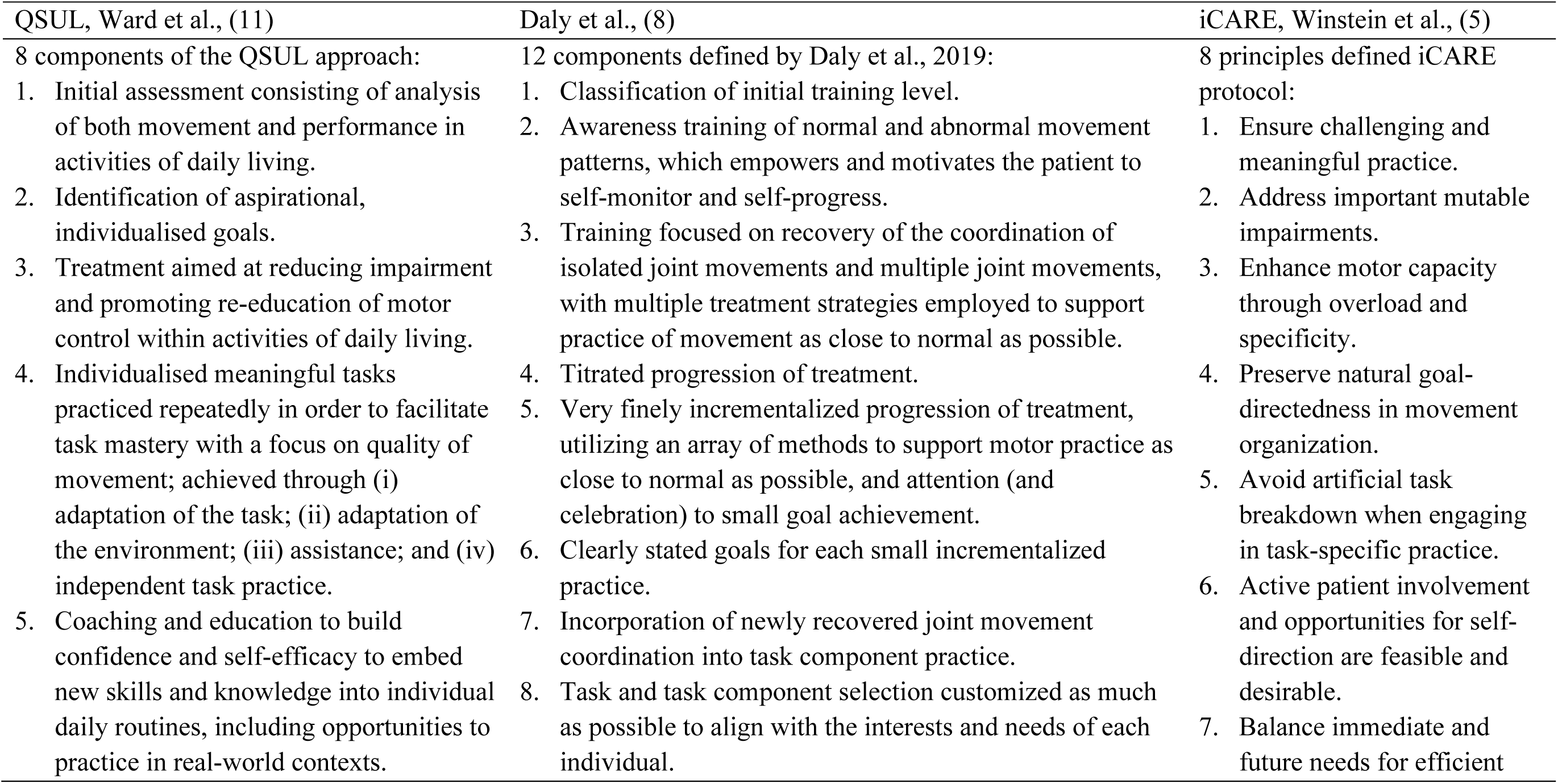
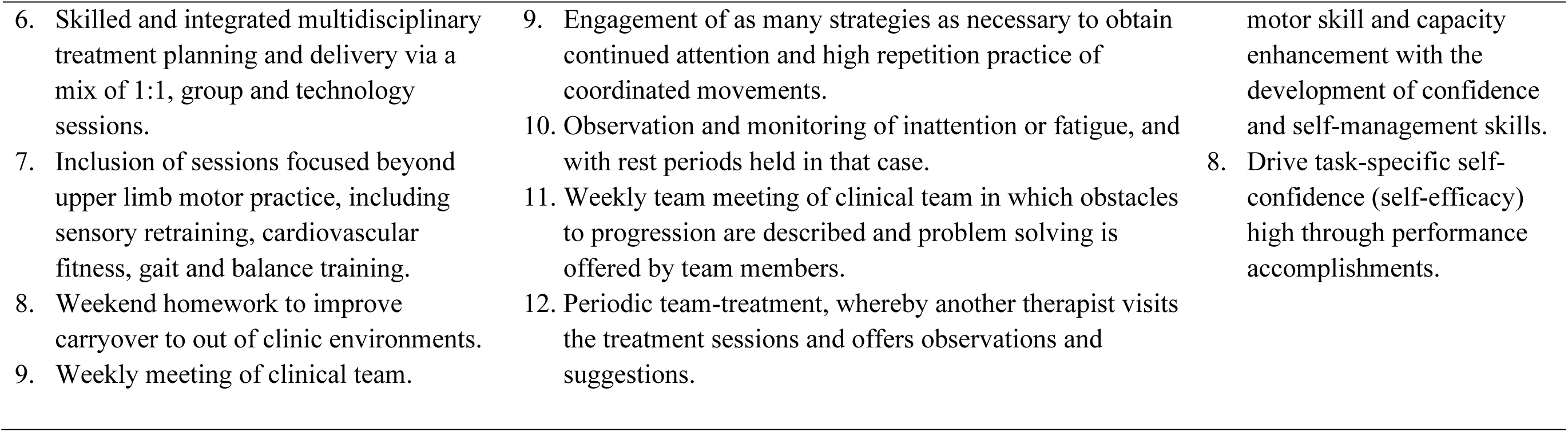
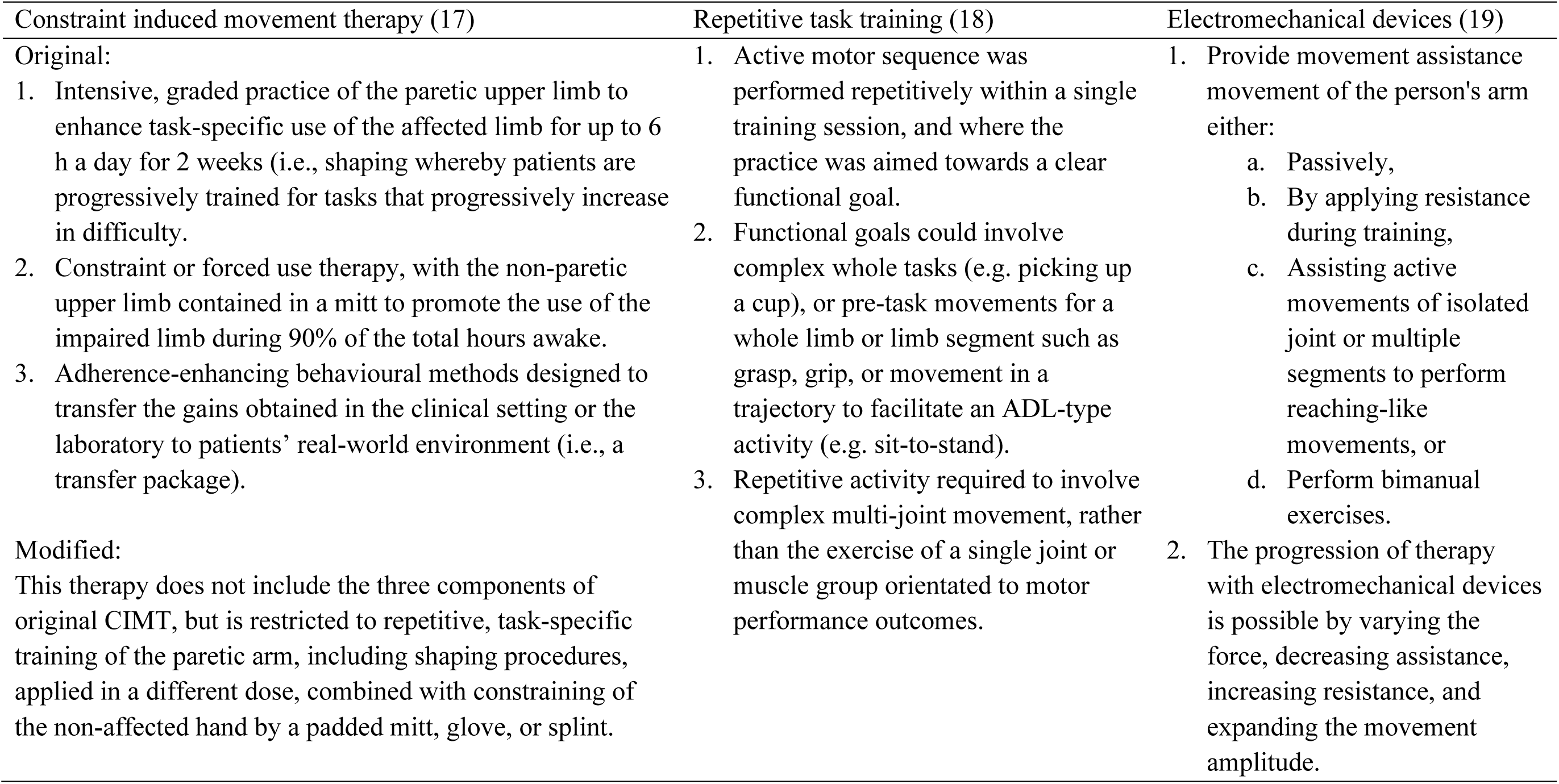
Components of upper limb stroke recovery treatments: (A) key components of interventions tested in individual studies and (B) components of interventions classes evaluated by systematic reviews.

### Strengths and limitations

Each focus group was facilitated by an independent person not involved in any aspect of the programme. Focus groups were independently transcribed by an external source. Gaining the perspective of multiple users enabled corroboration and triangulation of data and themes.

This study does however have a small sample size relative to the number of patients that have gone through the QSUL Programme (n>200) (11), as well as few male clinicians and limited carers in attendance at the focus groups. Collectively, these limitations do impact generalisability. Finally, focus groups were completed (for some) months after participation in the programme, therefore recall bias may be an issue.

## Conclusion

This study provides an interesting perspective of how an intensive upper limb neurorehabilitation programme is perceived by stroke survivors and clinicians involved. While the ‘gruelling’ intensity provided in this programme was considered important, creating individualised training opportunities that equip stroke survivors with skills, resources and knowledge to drive their own recovery for the longer term was equally emphasized. These considerations are important in determining the content of an upper limb neurorehabilitation trial that more accurately reflects effective clinical practice. Tools such as TIDieR (16) are key to defining the important components of a therapy programme and understanding the rationale for inclusion of each component and potential impact on recovery.

## Data Availability

Available on request by a qualified researcher.

## Author contributions

KSH, NW, KK, FB conceived the study. KSH, KK, FB, AS developed the focus group guide, thematically analysed the focus groups and wrote the manuscript; all authors intellectually contributed, reviewed and edited the manuscript; KSH led the focus groups, AS led participant recruitment.

## Patient and public involvement

Patients and public were participants but were not involved in the design of this study.

## Acknowledgements

Thanks to all the physiotherapists and occupational therapists at The National Hospital for Neurology and Neurosurgery, Queen Square, who have treated patients on this programme. Thanks to all the patients who have taken part in the programme. Thanks to UCLH Charities, Friends of UCLH and The National Brain Appeal for funding to purchase equipment used in the programme.

KSH received fellowship support from the National Health and Medical Research Council (NHMRC) of Australia (GNT1088449) and Michael Smith Foundation for Health Research British Columbia Canada (15980). KSH was also supported by a trainee travel award from the Canadian Partnership for Stroke Recovery, Future Leader Grant from the Stroke Foundation and Bayer Science and Education Foundation Award for Early Excellence in Science, which collectively supported her to undertake this collaborative research work. KSH is employed by The Florey Institute of Neuroscience and Mental Health, who acknowledges the strong support from the Victorian Government and in particular funding from the Operational Infrastructure Support Grant.

## Data availability

Data are available upon request to Dr’ s Ward or Hayward from credible researcher.

### Appendix 1

Please see the following from UCL https://protect-au.mimecast.com/s/T7N9Cp8AxKsVmrXpcPHQiV?domain=ethics.grad.ucl.ac.uk “Service Evaluation is undertaken to benefit those who use a particular service and is designed and conducted solely to define or judge current service. Your participants will normally be those who use the service or deliver it. It involves an intervention where there is no change to the standard service being delivered (e.g. no randomisation of service users into different groups). This does not require ethical approval.”

